# Utility of circulating tumor DNA for detection and monitoring of endometrial cancer recurrence and progression

**DOI:** 10.1101/2020.03.04.20030908

**Authors:** Esther L. Moss, Diviya N. Gorsia, Anna Collins, Pavandeep Sandhu, Nalini Foreman, Anu Gore, Joey Wood, Christopher Kent, Lee Silcock, David S. Guttery

## Abstract

Despite the increasing incidence of endometrial cancer (EC) worldwide and the poor overall survival of patients who recur, no reliable biomarker exists for detecting and monitoring EC recurrence and progression during routine follow-up. Circulating tumor DNA (ctDNA) is a sensitive method for monitoring cancer activity and stratifying patients that are likely to respond to therapy. As a pilot study, we investigated the utility of ctDNA for detecting and monitoring EC recurrence and progression in 13 patients using targeted next-generation sequencing (tNGS) and personalized ctDNA assays. Using tNGS, at least 1 somatic mutation at a variant allele frequency (VAF) >20% was detected in 69% (9/13) of patient tumors. The four patients with no detectable tumor mutations at >20% VAF were whole exome sequenced, with all four harboring mutations in genes not analyzed by tNGS. Analysis of matched and longitudinal plasma DNA revealed earlier detection of EC recurrence and progression and dynamic kinetics of ctDNA levels reflecting treatment response. We also detected acquired high microsatellite instability (MSI-H) in ctDNA from one patient whose primary tumor was MSI stable. Our study suggests that ctDNA analysis, and in particular MSI analysis in ctDNA could become a useful biomarker for early detection and monitoring of EC recurrence and progression.

## INTRODUCTION

Endometrial cancer (EC) is the second most common female cancer in the US, with over 60,000 women diagnosed each year [1]. In contrast to the high survival rate in patients with primary disease, the prognosis upon recurrence is poor, with a 5-year survival rate in patients with distant recurrence of 17% [2, 3], and the majority of EC recurrences in high-risk cases developing distant metastases within the first 3 years post-treatment [4]. There is no specific blood protein biomarker that is recommended for use following an EC diagnosis; instead, regular clinical examination is advised with imaging. The ovarian cancer markers CA125 and HE4 have been shown to significantly correlate with prognosis; however, their detection rates in metastatic disease are low, 54% and 75% respectively, and even lower with local recurrence, 39% and 16% [5]. Hence, more effective biomarkers are required that can detect EC recurrence and disease progression earlier, as well as reliably reflecting the underlying kinetics of the disease.

Cell-free DNA (cfDNA) can be extracted from plasma and the tumor-derived fraction of cfDNA, known as circulating tumor DNA (ctDNA) is rapidly becoming a quick and sensitive biomarker for tracking minimal residual disease and accurately reflecting therapeutic response in patients with actionable hotspot mutations [6, 7], chromosomal rearrangements [8], and gene amplifications [9, 10]. Although many studies have shown the ability of ctDNA for tracking and monitoring disease in cancers such as lung [11], breast [12] and colon cancer [13], few studies have highlighted its promise in EC. CtDNA has been detected in 18% of primary ECs using next-generation sequencing [14], as well as showing elevated levels 6 months before a rise in CA125 or radiological evidence of recurrence on CT imaging in gynecological cancers [15]. However, to date evidence is sparse highlighting the utility of ctDNA for detecting EC recurrence and monitoring treatment response during routine follow-up. Here, we used a combination of methods including targeted next-generation sequencing (tNGS), personalized ctDNA NGS analysis and digital droplet PCR (ddPCR) with the primary objective of detecting and monitoring EC recurrence and therapeutic response.

## METHODS

The study was approved by the Wales Research Ethics Committee 7 (17/WA/0342). All patients gave written informed consent prior to participation for use of their blood and tissue samples. Patients were recruited to the study if they were over the age of 18 and attending hospital follow-up appointments after an EC diagnosis, either after completion of primary treatment or after disease relapse. The reporting census date for this study was 15^th^ September 2019.

### Extraction and quantitation of DNA

Blood sampling and processing was performed as previously described [16]. Total cell-free DNA (cfDNA) was isolated from at least 3 mL of plasma using the QIAamp Circulating Nucleic Acid Kit (Qiagen) or using the MagMAX Cell-Free DNA Isolation Kit (Thermofisher) in combination with the Kingfisher Flex System (Thermofisher) in an automated manner according to manufacturer’s instructions. cfDNA levels (ng/mL) were converted to copies/mL plasma assuming 3.3 pg DNA per haploid genome. Isolation of DNA from lymphocytes and quantitation of total cfDNA was as described previously [16]. FFPE tumor DNA was extracted from 1.5 mm tissue cores using the Qiagen GeneRead Kit according to manufacturer’s instructions.

### Targeted next generation sequencing

Targeted NGS was performed on 20 ng of FFPE tumor DNA and lymphocyte DNA, and at least 5 ng of cfDNA (range 5 – 50 ng) using the Oncomine™ Pan-cancer cfDNA assay (Thermofisher – 52 genes covering >900 COSMIC mutations and 12 CNV regions). Libraries were prepared using the Ion Chef and sequenced on Ion 540 chips using the Ion S5 XL according to manufacturer’s instructions to a minimum average read depth of at least 10,000x. Sequencing data was accessed through the Torrent Suite v5.6 and analyzed using Ion Reporter v5.6. For FFPE and lymphocyte DNA, the Oncomine TagSeq Pancan Tumor w1 workflow was used. For cfDNA, the Oncomine TagSeq Pancan Liquid Biopsy w1 workflow was used. Initially, targeted NGS using the Oncomine™ Pan-Cancer cfDNA assay was conducted to identify the somatic mutations present in the primary tumor tissue in all cases; in two cases additional biopsies from metastatic and recurrent disease were also sequenced.

### Digital droplet PCR

Droplet digital PCR (ddPCR) was performed to analyse the *TP53* p.Y220C mutation (assay numbers dHsaCP2500536 and dHsaCP2500537; Bio-Rad Laboratories) and *KRAS* p.G12D mutation (assay numbers dHSaCP2000001 and dHsaCP2000002) according to manufacturers’ instructions. Thermal cycling conditions were: 10-minute hold at 95 °C, 40 cycles of 95 °C for 15 seconds and then 55 °C for 60 seconds. Raw fluorescence amplitude was analyzed using the Quantasoft version 1.6.6.0320 software (Bio-Rad Laboratories) [17].

### Whole exome sequencing and personalized ctDNA sequencing

The Cell3 Target Whole exome kit (Nonacus Ltd, Birmingham) and Illumina NextSeq500 was used to perform whole-exome sequencing on 150-200 ng of tumor DNA from each FFPE primary tumor block and 50ng of germline DNA at an average read depth of 200x for tumor DNA and 50x for matched lymphocyte samples. Personalized ctDNA panels were developed for analysis of at least 2 SNVs per patient (Table S1) using at least 5 ng of cfDNA (range 16 – 25 ng). Libraries were prepared using the Cell3^(tm)^ Target Custom Panel (Nonacus Ltd, Birmingham, UK) and sequenced using an Illumina MiSeq to a minimum depth of 30,000x.

### Bioinformatic analysis

BAM files from whole exome sequencing or personalized ctDNA assays were prepared using an in-house pipeline to process sequencing output after demultiplexing. The BAM files were then aligned to human genome reference 38 (GRCh38). Somatic variants arising from WES were called using two variant callers, Platypus [18] and Mutect2 [19] whilst variants detected in cfDNA using the personalized cfDNA assay were called using solely Platypus. Variants were called using a minimum base quality and minimum mapping quality of 20, with a minimum of 1 read fitting these parameters for variants to be identified. Additionally, variants were filtered for those with an 11-base window around to the variant location to check for variants with a minimum quality < 15. The output VCF files were annotated using ANNOVAR [20] to identify and annotate the subsets and variants. Microsatellite instability was analyzed using MSIsensor [21] for detection of mononucleotide repeats at several genomic sites (BAT-25, BAT-26, NR-21, NR-24 and NR-27) in tumor alleles as determined by a shift in ≥ 3bp in each marker. MSI-high tumors were defined as those with ≥ 2 unstable markers, whilst samples with 1 unstable markers were classified as MSI low tumors and those with 0 unstable markers were classified as microsatellite stable (MSS).

## RESULTS

In total, a cohort of 13 cases of mixed risk and mixed histology were analyzed. At the reporting census date, six patients had recurred/progressed, three had died (two of their disease and one of a non-EC cause) and four patients were clinically well with no clinical or radiological sign of recurrence/progression (Table 1).

**Table 1:** Tumor characteristics and outcome of study cohort (n=13)

| Patient | Histology | Stage at diagnosis | Recurrence | Site of recurrence | Outcome |
| --- | --- | --- | --- | --- | --- |
| 1 | Endometrioid G1 | IB | Yes | Pelvic | Alive with disease |
| 2 | Endometrioid G3 | IB | Yes | Distant | Died with disease |
| 3 | Serous | IA | Yes | Distant | Alive with disease |
| 4 | Endometrioid G1 | IB | Yes | Pelvic, distant | Alive with disease |
| 5 | Endometrioid G3 | IVB | Yes | Distant | Alive with disease |
| 6 | Endometrioid G2 | IVB | Yes | Distant | Died with disease |
| 7 | Endometrioid G1 | IVB | Yes | Distant | Alive with disease |
| 8 | Carcinosarcoma +<br>Serous | IVB | Yes | Pelvic, distant | Alive with disease |
| 9 | Endometrioid G1 | IA | Yes | Vaginal vault | Alive no disease |
| 10 | Endometrioid G1 | I | No | NA | Died no disease |
| 11 | Serous | IIIC1 | No | NA | Alive no disease |
| 12 | Clear Cell | IIIB | No | NA | Alive no disease |
| 13 | Carcinosarcoma | IIIA | No | NA | Alive no disease |

### Sequencing of primary tumors

Initially, targeted NGS using the Oncomine™ Pan-Cancer cfDNA assay was conducted to identify the somatic mutations present in the primary tumor tissue in all cases; in two cases additional biopsies from metastatic and recurrent disease were also sequenced for cases 5 and 9 respectively. At least 1 somatic hotspot mutation at an arbitrary cut-off of ≥ 20% VAF was detected in 9/13 (69%) patient tumors that was not present in the matched lymphocytes (Table S2). In the other four cases, matched tumor and normal samples were sequenced using a whole exome (WES) approach, then used to guide development of a custom targeted cfDNA panel (Table S1). For details of the mutations detected in each patient and selected for custom ctDNA panel design, see Table S2. Two of the patients sequenced using WES (patients 1 and 2) were found to have an unusually high number of single nucleotide variants (SNVs) with 301 SNVs and 131 SNVs in 37MB respectively; using MSIsensor, patient 1 was identified as MSI-H (5/5 markers positive) and patient 2 harboured a *POLE* p.G541E mutation of unknown significance, but her primary tumor was MSS.

### ctDNA can detect EC recurrence and progression earlier than scans

The median lead time of ctDNA over radiological imaging or clinical recurrence in patients for whom we had sample available prior to recurrence or progression was 2.5 months (n = 6, range 1 – 8 months) (Figure 1). ctDNA was detected in all four patients (100%) who were diagnosed with stage I disease at diagnosis and who subsequently recurred (Figure 1, Table S2). For patient 1 the lead time of ctDNA over radiological imaging was 8 months and there was 16 months between the first positive ctDNA and the patient developing symptoms (Figure 1). Two of the stage I patients (cases 1 and 2) had serial samples analyzed and disease progression was confirmed with increasing ctDNA levels in concordance with both clinical and imaging progression (case 2). ctDNA was detected in the four patients (100%) who were diagnosed with stage IV disease and had progressive disease on imaging (Figure 1, Table S2).

**Figure 1:**
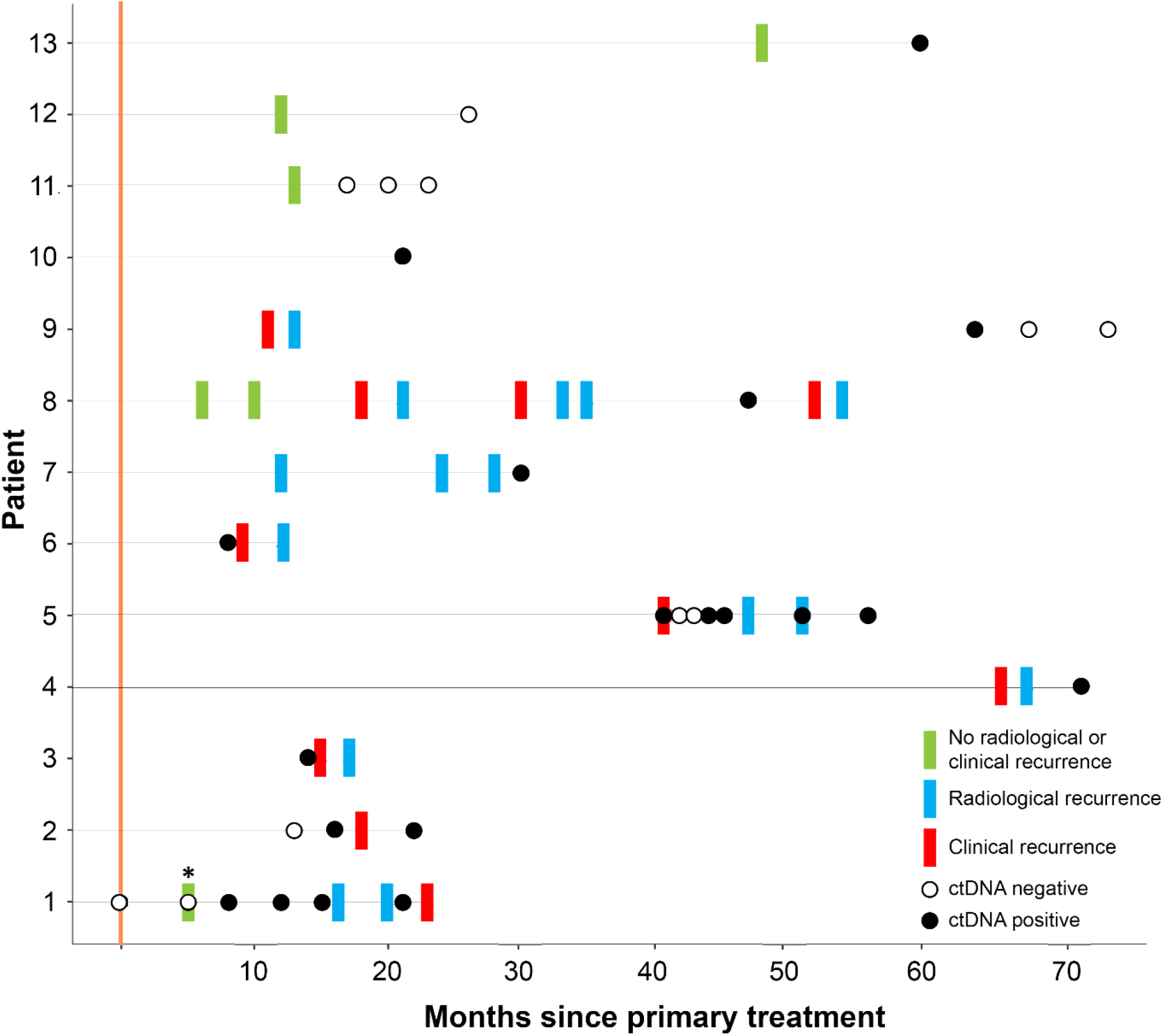
Summary of each patient’s results of clinical imaging and results of serial samples in terms of detection of ctDNA. Asterisks above sample for patient 1 indicates mutation was present in IGV but not called due to high strand bias.

### ctDNA accurately reflects EC disease kinetics during treatment

ctDNA levels accurately mirrored the radiological response of a patient undergoing chemotherapy treatment for recurrent EC, with a clonal *TP53* p.Y220C mutation becoming undetectable during treatment, before rising again prior to radiological progression (Figure 2). An additional two patients (cases 9 and 11) diagnosed with stage I EC received radiotherapy, one for a vaginal vault recurrence and the other as primary treatment, respectively. For case 11, ctDNA was not detected in three plasma samples following primary radical radiotherapy, in keeping with clinical findings of complete response (Figure 1). ctDNA was detected following treatment of the vault recurrence (case 9 – Table S2); however, became negative with time and remained negative, in keeping with clinical and radiological findings of complete resolution of the recurrence and no ongoing disease.

**Figure 2:**
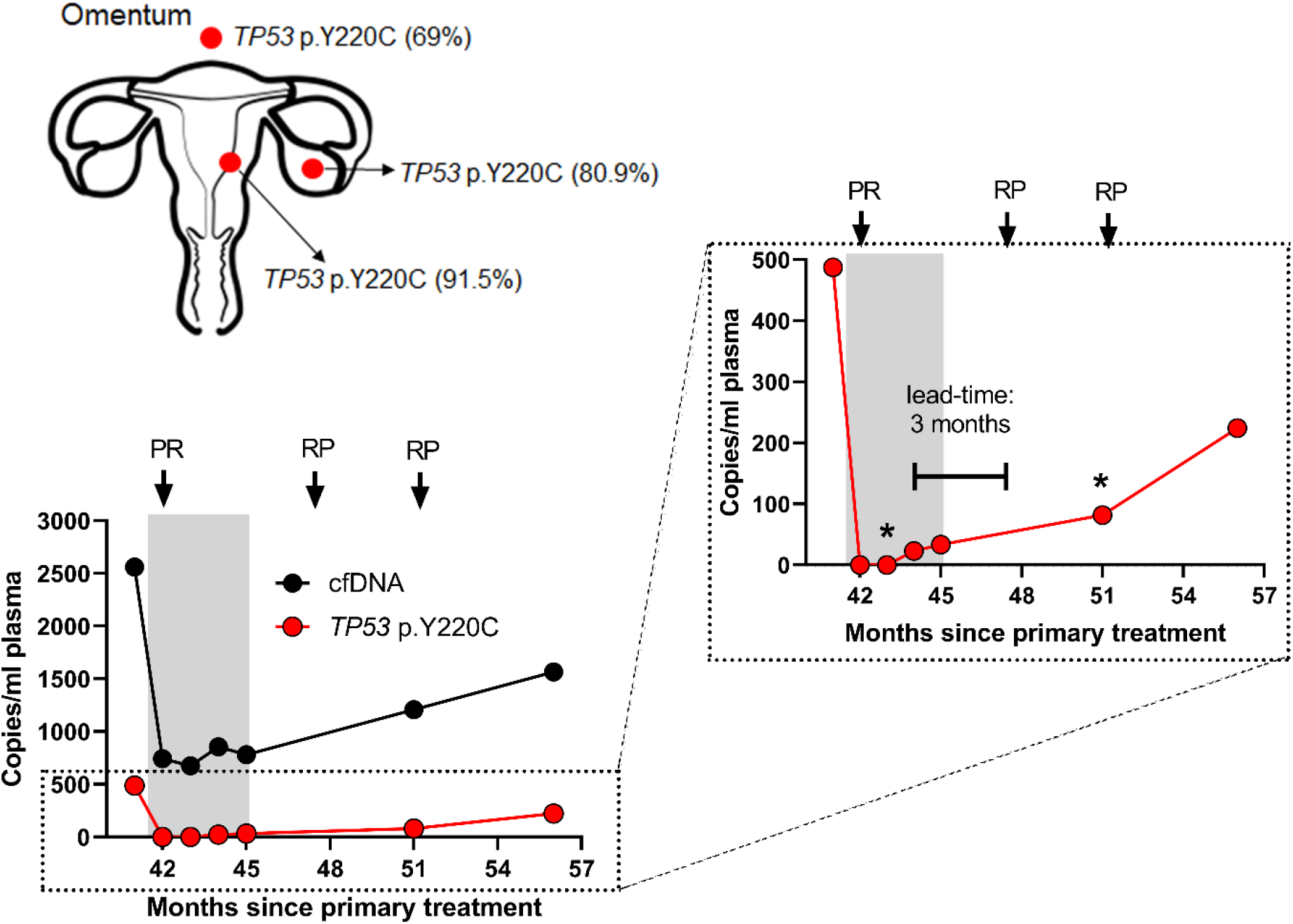
Multiregional sampling from patient 5 alongside the respective variant allele frequencies of the major driver mutation, *TP53* p.Y220C, identified in the primary tumor sample (located in the endometrium), local metastatic lesion (located in the ovary) and the distal metastatic lesion (located in the omentum). The levels of cfDNA and ctDNA in pre-treatment and post-treatment plasma samples are shown in the accompanying left-hand graph, clearly demonstrating a rise in ctDNA levels 3 months prior to radiological relapse (right-hand). *samples analyzed exclusively by ddPCR. Grey shaded area indicates treatment with carboplatin and paclitaxel. PR = Partial response; RP = radiological progression.

### Follow-up of high-risk EC cases

In the three cases with high-risk EC at diagnosis (stage III non-endometrioid histology) ctDNA as detected in only one case (Patient 13 - Figure 1, Table S2). All patients were alive at the consensus date with no clinical or radiological sign of recurrence and continue to be followed-up.

### Longitudinal ctDNA can detect acquired microsatellite instability

Only one patient’s tumor (case 1) was classified as MSI-H with all five markers displaying microsatellite instability, which was confirmed using ctDNA (3/5 markers positive – BAT25, NR-24 and NR-27) (Figure 3A). In addition, ctDNA was able to identify the acquisition of microsatellite instability in a case (case 2) whose primary tumor was microsatellite stable (MSS). The patient had received adjuvant chemotherapy but had subsequently recurred. Longitudinal samples revealed the acquisition of MSI-H status, with two of the five markers (NR-24 and NR-27) displaying MSI in the last plasma sample (Figure 3B, Table S2).

**Figure 3:**
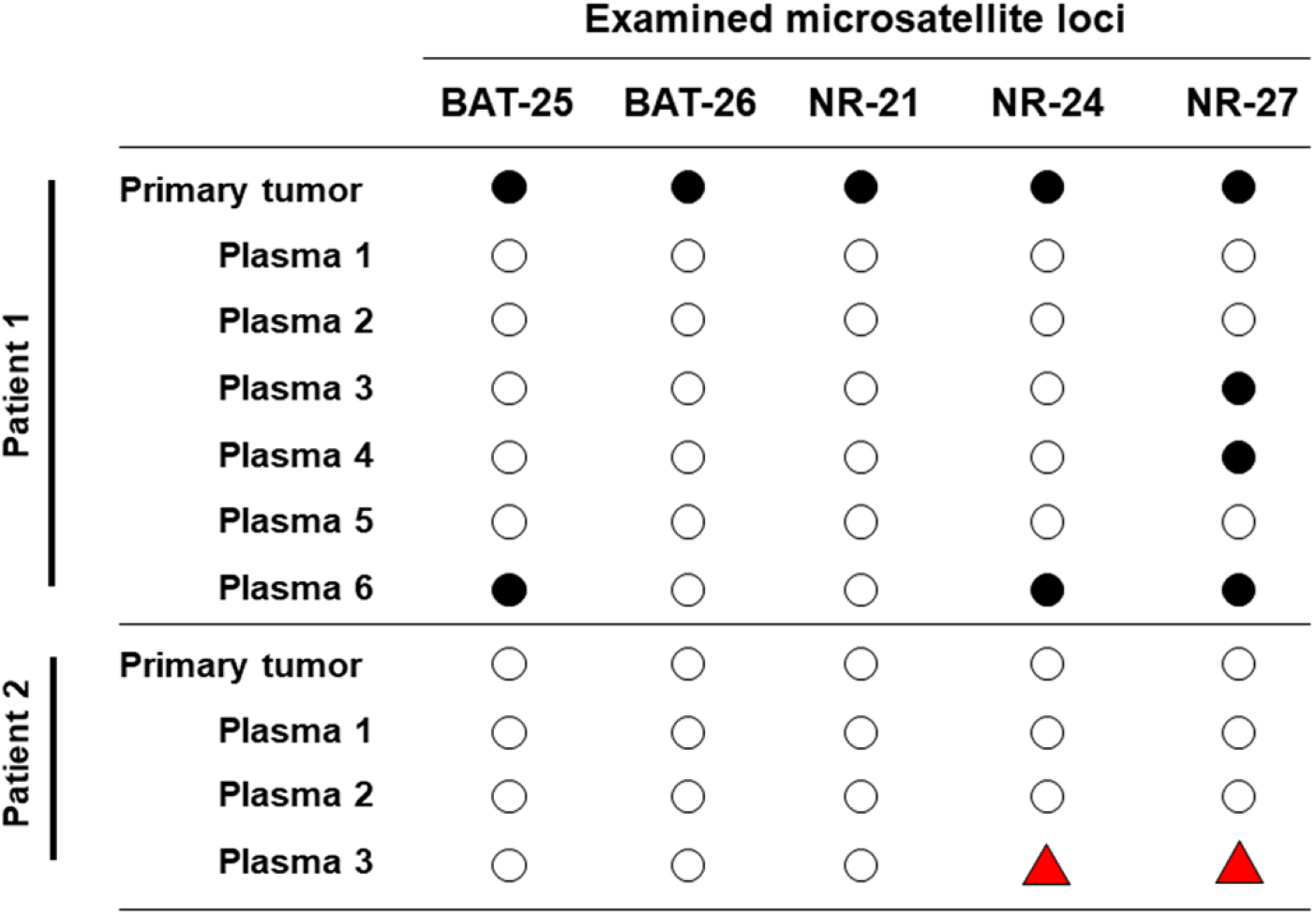
Microsatellite status in the primary tumors and longitudinal plasma samples of patients 1. (MSI-H) and 2 (MSS). ○ = alleilic shift not detected, ● = alleilic shift detected, 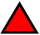 = alleilic shift has changed from negative to positive.

### ctDNA is reflective of EC tumor heterogeneity and evolution

Analysis of the primary, metastatic and recurrent tumor biopsies and matched plasma DNA for two cases (cases 5 and 9) demonstrated that mutations identified in biopsies from different locations were present in the ctDNA but that additional mutations could also be identified. Truncal *TP53* p.Y220C (case 5) and *PIK3CA* p.H1047R (case 9) mutations were identified in primary and recurrent tumors but additional mutations, *MET* p.T1010I, *FGFR2* p.S252W and *KRAS* p.G12A, were detected in the ctDNA at recurrence that were not present in the tumor samples (Table S2).

## DISCUSSION

We have shown that ctDNA is highly sensitive in identifying endometrial cancer recurrence in different histological subtypes and grades, and have a lead time of up to 16 months over patient reported symptoms and up to 8 months compared to imaging. In addition, we have shown that ctDNA levels accurately reflect disease activity, with earlier identification of response/progression than on CT imaging. We also report the first case of ctDNA identifying the acquisition of microsatellite instability in a case of EC recurrence, an increasingly important stratifying feature for determining optimum patient treatment.

Our results are in keeping with studies published in other tumor types and supports the clinical utility of ctDNA in early detection of recurrence in endometrial cancer [15] and other solid tumors [12, 13]. The use of different technologies for ctDNA detection has given further insight into the optimum methods that can be taken forward for use in a clinical trial determining the potential for this new development on patient management. We have shown a benefit in using WES to profile the tumor tissue and tracking mutations with a customised targeted cfDNA panel relative to using a driver gene approach, which did not identify any mutations in four tumor samples, equating to approximately one third of our cohort who otherwise would not have been monitored. Additionally, the customised targeted panel can give greater informative value in monitoring of tumor evolution, identifying somatic variants not present in the primary tumor tissue. This further highlights the beneficial value of using a targeted customised panel approach over a driver gene approach as it may not elucidate the evolving genetic proportions as accurately as the personalized approach.

At present there is no EC specific blood tumor marker to monitor for disease recurrence, unlike CA125 and ovarian cancer. Our results have shown the potential of ctDNA to monitor patients undergoing treatment, as compared to waiting for radiological response. Parkinson et al., reported that chemotherapy response for the treatment of high-grade serous ovarian cancer was detected earlier with ctDNA, as compared to CA125, with a nadir of 37 days compared to 84 days respectively, and ctDNA was more prognostic than CT imaging [22]. Resistance to first line chemotherapy for EC is known to be a greater issue than with ovarian cancer [23], and should our results be replicated in a larger population it opens up the potential for ctDNA identification of non-efficacious treatments at an earlier time point, rather than waiting for imaging assessment of disease response.

Our results have also shown an additional benefit of longitudinal monitoring of patients using personalized assays containing MSI markers with the discovery of acquired MSI-H status through serial plasma sampling. ctDNA analysis has been shown to detect MSI [24], reflect tumor heterogeneity [25] and to track tumor evolution [11]. Acquired MSI-H status has previously been shown in prostate cancer [26] however, to our knowledge, this is the first report of acquired MSI-H status in EC. The presence of a MMR gene deficiency is known to be positively associated with response to PD-1 inhibitors [27] and the approval of Pembroluzumab has the potential to benefit up to 30% of advanced EC patients [28, 29]. This has led to the recommendation that all EC patients undergoing MSI testing [30]. Indeed, in the relapse setting, the use of Pembrolizumab has been modeled to be cost-effective in MSI-H patients, as compared to single agent pegylated liposomal doxorubicin or bevacizumab [31]. The identification of acquired MSI mutations in relapsed EC highlights another population of patients who could potentially benefit from a PD-1 inhibitor but who would not be eligible if their MSI status was determined from their primary tumor alone. The ability of ctDNA to detect such tumor evolutions through a test with high patient acceptability [32] and without the need for invasive biopsies or surgery could further support the development of personalized patient management and hopefully improved patient outcomes.

In conclusion, ctDNA monitoring is able to accurately monitor the activity of endometrial cancer and diagnose recurrence. Further work is needed to determine the lead time and its ability to identify local, as well as distant, recurrent disease

## Data Availability

All whole exome sequencing data will be deposited in the European Genome Phenome Archive on publication.

## Author contribution

E.L.M. and D.S.G designed the study, E.L.M., D.S.G., L.S. and D.N.G performed experiments, analyzed data and wrote the manuscript; E.L.M., A.C., P.S., N.F., A.G., J.W. and C.K. conducted informed consents and collected samples and clinical data; E.L.M and D.S.G supervised the study.

